# EFFECT OF MALARIA ON THE RETICULOCYTE COUNT OF FEMALE STUDENTS OF A UNIVERSITY IN ELELE, RIVERS STATE

**DOI:** 10.1101/2025.05.25.25328311

**Authors:** C. C. Echiejini, A. I. Ezeamalu, Q-M Muoghara, S. E. Idimogu, R. A. Ikpeama, B. J. Okonko, I. O. Okonko

## Abstract

**Background:** Malaria remains a significant public health challenge in sub-Saharan Africa, with profound haematological consequences, particularly anaemia.

**Objective:** This study investigated the impact of *Plasmodium falciparum* infection on reticulocyte counts among female undergraduate students at Madonna University, Nigeria, a hyperendemic region.

**Methods:** A cross-sectional analysis of 40 participants aged 15-30 years was conducted during peak transmission season (April-October). Malaria diagnosis was confirmed through Giemsa-stained microscopy and rapid diagnostic testing, while reticulocyte counts were performed manually using new methylene blue staining.

**Results:** The result revealed 80% malaria prevalence, with the highest rates in 18-20 year-olds (50%). Parasite density distribution showed 90.6% low, 6.3% moderate, and 3.1% high parasitemia cases. Reticulocyte counts were significantly lower in high parasitemia cases (0.8%) compared to low/moderate parasitemia (1.4%) and uninfected students (1.3%). Age-related patterns emerged, with the lowest reticulocyte counts in infected 15-17 year-olds (0.36%) and highest in ≥24 year-olds (1.67%).

**Conclusion:** The findings demonstrate malaria’s hematological impact in this population and suggest the utility of reticulocyte monitoring in assessing infection severity.

## 1. Introduction

Malaria remains one of the most devastating parasitic diseases worldwide, with an estimated 247 million cases and 619,000 deaths reported in 2021, predominantly in sub-Saharan Africa [1]. The disease is caused by *Plasmodium* parasites, transmitted through the bites of infected female *Anopheles* mosquitoes, and poses a significant public health burden due to its association with anemia, impaired cognitive function, and economic losses in endemic regions [2]. Among the most affected populations are young adults, particularly university students in tropical regions, where environmental conditions, limited access to preventive measures, and communal living exacerbate exposure risks [3].

A critical yet understudied aspect of malaria infection is its impact on erythropoiesis—the process of red blood cell (RBC) production. Malaria-induced hemolysis and dysregulated bone marrow function often lead to anemia, a common complication that contributes to fatigue, reduced academic performance, and long-term health consequences [4]. Reticulocytes, immature RBCs containing residual ribosomal RNA, serve as a key biomarker of bone marrow activity. In healthy individuals, reticulocyte counts typically range between 0.5% and 2.5% of total RBCs, reflecting normal erythropoietic function [5]. However, during malaria infection, reticulocyte dynamics may shift dramatically. Some studies report increased reticulocyte production as a compensatory mechanism for parasite-induced RBC destruction,[6] while others document suppressed reticulocytosis due to inflammation-mediated bone marrow suppression [7].

The relationship between malaria and reticulocyte response remains poorly characterized in young adult populations, particularly in university settings where stress, nutritional factors, and high transmission rates may influence hematological outcomes [8]. Existing research has primarily focused on children and pregnant women, who are considered high-risk groups [9]. However, university students—especially those in endemic regions—represent a vulnerable yet overlooked demographic. Frequent exposure to malaria, combined with inadequate healthcare access and irregular use of preventive measures, may predispose them to chronic anemia and its associated complications [10].

Madonna University, located in Elele, Nigeria, is situated in a high malaria transmission zone, making it an ideal setting to investigate these hematological changes [11]. A prior study found an 80% malaria prevalence among female students at this institution, with varying degrees of parasitemia [12]. However, no study has yet examined how these infections affect reticulocyte counts, which could provide critical insights into the severity of hemolytic anemia and bone marrow response in this population.

Given the high malaria burden among Nigerian university students and the potential academic and health implications of malaria-induced anemia, this study aims to compare reticulocyte counts between malaria-positive and malaria-negative female students at Madonna University, assess the correlation between malaria parasite density and reticulocyte levels to determine if disease severity influences erythropoietic response and evaluate age-related variations in reticulocyte production to identify potential demographic risk factors for malarial anemia.

This research will contribute to the limited body of knowledge on malaria-related hematological changes in young adults and provide evidence to support targeted interventions, such as routine anemia screening and improved malaria prophylaxis programs on university campuses. Additionally, the findings may aid in identifying subclinical hemolysis in asymptomatic carriers, a critical step toward reducing the long-term health consequences of recurrent malaria infections in endemic regions.

## 2. Materials and Methods

### 2.1. Study Design and Setting

This study employed a cross-sectional analytical design to evaluate the impact of malaria infection on reticulocyte counts among female students at Madonna University, Elele Campus, Rivers State, Nigeria. The university is situated in a region characterized by hyperendemic malaria transmission with year-round transmission intensity [13]. Data collection was conducted during the peak malaria transmission season (April to October 2023) to maximize the detection of active infections and ensure robust representation of malaria-related hematological changes.

### 2.2. Study Population and Sampling

The study population consisted of female undergraduate students aged 15 to 30 years, selected through stratified random sampling across all academic levels to ensure representativeness. The sample size was calculated using Leslie Kish’s formula [14], which is appropriate for prevalence studies. The calculation incorporated a baseline malaria prevalence of 56.3% [15], derived from previous epidemiological studies conducted at the university, with a 95% confidence interval and 5% margin of error. After accounting for a 10% attrition rate, the final sample size was determined to be 40 participants.

Participants were included based on specific criteria to control for confounding variables. Eligible students were required to have resided on campus for at least six months prior to the study, ensuring consistent exposure to local malaria transmission patterns. Individuals who had taken antimalarial medications within the preceding four weeks were excluded to avoid pharmacological interference with hematological parameters. Additionally, students with known hematological disorders or those who had received blood transfusions in the recent past were excluded to maintain the integrity of reticulocyte measurements. Also, severely malnourished individuals (BMI <18.5 kg/m^2^) [16] were excluded.

### 2.3. Data Collection Procedures

#### 2.3.1. Malaria Diagnosis

Venous blood samples (3 mL) were collected from each participant using standard phlebotomy techniques and immediately transferred into EDTA-coated tubes to prevent coagulation. Samples were processed within two hours of collection to minimize pre-analytical variability. Malaria diagnosis was performed using two complementary methods to ensure diagnostic accuracy.

Giemsa-stained thick and thin blood films [17] were prepared and examined under light microscopy by experienced laboratory technicians. Thick films were used for initial parasite detection due to their high sensitivity (capable of detecting 5–10 parasites per microliter of blood), while thin films facilitated *Plasmodium species* identification. Parasite density was quantified according to World Health Organization guidelines [18] and categorized as follows: + (1–100 parasites per 100 fields), ++ (101–1,000 parasites per 100 fields), or +++ (>1,000 parasites per 100 fields).

In parallel, rapid diagnostic testing (SD BIOLINE Malaria Ag Pf/Pan®) [19] was performed to detect *Plasmodium falciparum*-specific histidine-rich protein-2 (HRP-2) and pan-malarial lactate dehydrogenase (pLDH) antigens. This dual diagnostic approach enhanced the sensitivity and specificity of malaria detection, particularly in cases with low parasitemia.

#### 2.3.2. Reticulocyte Counting

Reticulocyte enumeration was conducted using a standardized manual counting technique with new methylene blue stain [20]. Briefly, equal volumes of blood and 1% new methylene blue solution were mixed and incubated at room temperature for 15 minutes to allow for sufficient staining of ribosomal RNA within reticulocytes. Smears were then prepared and examined under oil immersion microscopy (1000× magnification).

A minimum of 1,000 red blood cells were counted per sample, and reticulocytes were expressed as a percentage of the total erythrocyte population. To account for variations in hematocrit levels among participants, the reticulocyte production index (RPI) was calculated using the formula: RPI = (Reticulocyte percentage × Patient’s hematocrit) / (45 × Maturation time) [21]. This adjustment provided a more accurate reflection of bone marrow erythropoietic activity, particularly in anemic individuals.

### 2.4. Statistical Analysis

Data analysis was performed using SPSS version 25 (IBM Corp.). Descriptive statistics, including means, and frequencies, were computed to summarize participant characteristics and laboratory findings. Categorical variables were analyzed using chi-square tests. A p-value of <0.05 was considered statistically significant, with all tests conducted at 95% confidence intervals [22].

### 2.4. Ethical Considerations

The study protocol received ethical approval from the Madonna University Health Research Ethics Committee (Approval No: MUHREC/2023/041). Written informed consent was obtained from all participants after a detailed explanation of the study objectives, procedures, and potential risks. Confidentiality was maintained through anonymized data collection and secure storage of records. Students who tested positive for malaria were promptly treated with artemether-lumefantrine combination therapy following Nigeria’s national malaria treatment guidelines [23], but after sample collection for the experiment.

## 3. Results

Figure 1 shows that 32 students, representing 80% of the sample, tested positive for malaria parasites. Conversely, 8 students, making up 20%, tested negative.

**Figure 1:**
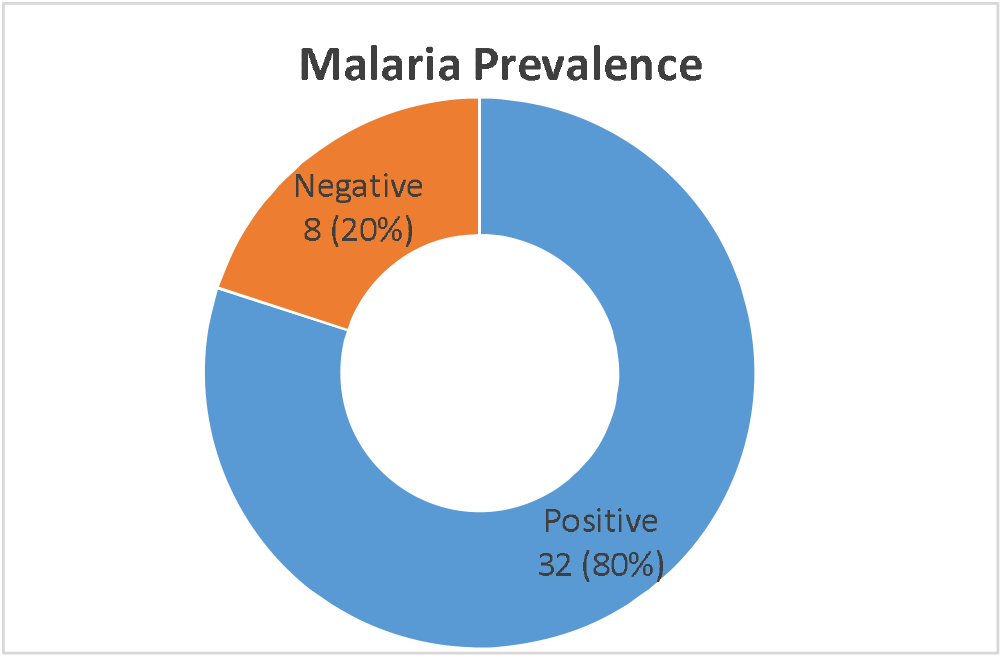
Malaria Prevalence.

**Table 1** shows the distribution of malaria parasite among age groups. It indicates that the highest presence of malaria parasites is in the 18-20 age group, with 50% of students testing positive, followed by the 21-23 age group at 34.375%. The 15-17 and 24 and above age groups have lower prevalence rates at 6.25% and 9.375%, respectively. Despite these variations, the chi-square test result (X^2^=5.399, p = 0.145) shows that the differences are not statistically significant (P > 0.05).

**Table 1.** Malaria Parasite Distribution by Age Group.

| Age Group (years) | Malaria Positive n (%) | Malaria Negative n (%) | Total |
| --- | --- | --- | --- |
| 15-17 | 2 (6.3) | 1 (12.5) | 3 |
| 18-20 | 16 (50.0) | 7 (87.5) | 23 |
| 21-23 | 11 (34.4) | 0 (0.0) | 11 |
| ≥24 | 3 (9.4) | 0 (0.0) | 3 |
| Total | 32 (100) | 8 (100) | 40 |

( $X^2 = 5.399$ , $p = 0.145$ ) $P > 0.05$ (Not significant)

Table 2 shows the distribution of malaria quantification across different age groups among students. Most students have low levels of malaria parasites, with 2 students in the 15-17 age group, 14 in the 18-20 age group, 10 in the 21-23 age group, and 3 in the 24 and above age group. Moderate levels of infection are only observed in 2 students within the 18-20 age group, while a high level of infection is noted in just 1 student from the 21-23 age group. The chi-square test (X2=3.975, p = 0.680) indicates that these differences are not statistically significant (P > 0.05), Parasite density grading showed 29 cases (90.6% of positives) with low parasitemia (+), 2 cases (6.3%) with moderate parasitemia (++), and 1 case (3.1%) with high parasitemia (+++).

**Table 2:** Malaria Density Distribution based on Age Group.

| Malaria quantification | Age Groups (years) |  |  |  |
| --- | --- | --- | --- | --- |
|  | 15-17 | 18-20 | 21-23 | 24 and above |
| + | 2 | 14 | 10 | 3 |
| ++ | - | 2 | - | - |
| +++ | - | - | 1 | - |

( $X^2 = 3.975$ , $p = 0.680$ ) $P > 0.05$ (Not significant)

Table 3 shows the mean distribution of reticulocyte count among female students in Madonna University based on age across different age groups. For the 15-17 age group, the 25 mean reticulocyte count is 0.36 in those with malaria and 1.20 in those without. In the 18-20 age group, the mean count is higher at 1.45 for those with malaria, slightly decreasing to 1.29 for those without. Students aged 21-23 with malaria have a mean reticulocyte count of 1.29, with no available data for those without malaria. Similarly, for students aged 24 and above, the mean count is 1.67 for those with malaria, with no data for those without.

**Table 3.**
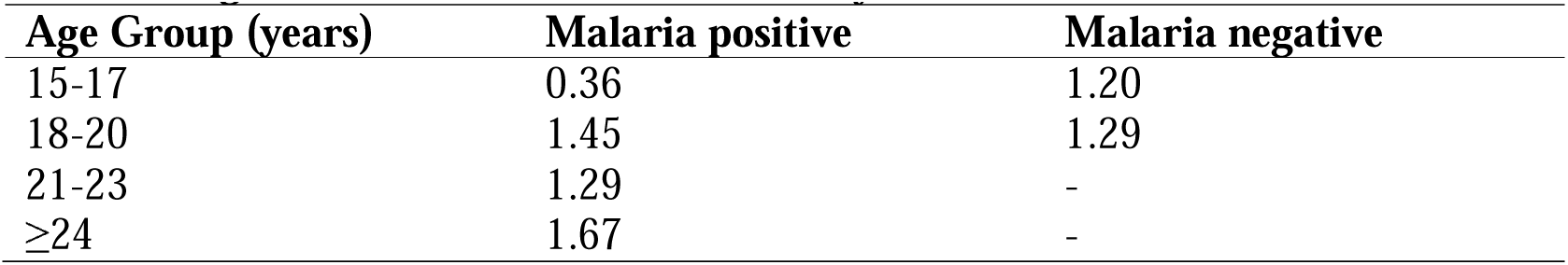
Age Distribution of Mean Reticulocyte Counts.

| Age Group (years) | Malaria positive | Malaria negative |
| --- | --- | --- |
| 15-17 | 0.36 | 1.20 |
| 18-20 | 1.45 | 1.29 |
| 21-23 | 1.29 | - |
| ≥24 | 1.67 | - |

Table 4 shows the average distribution of reticulocyte count among female students of Madonna University based on malaria quantification Students with low (+) and moderate (++) malaria quantification have an identical mean reticulocyte count of 1.4. Those with high (+++) malaria quantification have a lower mean count of 0.8. In comparison, students who tested negative for malaria have a mean reticulocyte count of 1.3.

**Table 4:** Average Distribution Of Reticulocyte Count Among Female Students Of Madonna University Based On Malaria Quantification.

| Malaria Quantification | Mean Reticulocyte Count |
| --- | --- |
| + | 1.4 |
| ++ | 1.4 |
| +++ | 0.8 |
| Negative | 1.3 |

## 4. Discussion

The findings of this study reveal important patterns in malaria prevalence and its hematological effects among female university students in an endemic region. The high malaria prevalence rate of 80% substantially exceeds Nigeria’s national average of 27% [24], likely reflecting both the university’s location in a high-transmission zone and the unique exposure risks associated with student living conditions. This elevated prevalence aligns with previous reports documenting similar infection rates among Nigerian university populations [25].

The age distribution analysis showed interesting though non-significant patterns (X^2^=5.399, p=0.145), with the highest infection rates occurring in the 18-20 year age group (50%) and the lowest in both the youngest (15-17 years, 6.3%) and oldest (≥24 years, 9.4%) participants. This bimodal distribution may reflect behavioral factors in the 18-20 year age groups, such as increased outdoor activities or reduced use of protective measures, though the lack of statistical significance suggests these differences may represent random variation rather than true age-dependent susceptibility [26].

The reticulocyte count analysis yielded several clinically relevant observations. Most notably, students with high parasitemia (+++) showed markedly lower mean reticulocyte counts (0.8%) compared to those with low/moderate parasitemia (1.4%) and uninfected students (1.3%). This finding provides clinical evidence supporting the concept of malaria-induced erythropoietic suppression in cases of high parasitic burden [27]. The similar reticulocyte counts between low and moderate parasitemia groups suggests this suppression may only occur beyond a certain threshold of parasite density.

Age-related patterns in reticulocyte response were particularly noteworthy. While the 15-17 year group showed the lowest reticulocyte counts among infected students (0.36%), the ≥24 year group demonstrated the highest counts (1.67%), potentially indicating more effective erythropoietic compensation in older students. The unexpectedly low reticulocyte counts in malaria-negative participants (mean 1.3%) compared to standard reference ranges (1.5-2.5%) [28] may reflect subclinical effects of previous infections or nutritional factors common in student populations [29].

## 5. Conclusion

This study demonstrates that malaria infection remains highly prevalent among university students in endemic regions, with significant effects on erythropoietic function. The key findings indicate that while malaria prevalence does not vary significantly by age in this population, hematological responses show important variations, particularly the marked reticulocyte suppression in high parasitemia cases. These results highlight the need for enhanced malaria prevention measures in university settings and suggest that reticulocyte monitoring may be valuable for assessing hematological impact in infected individuals.

## Data Availability

All data produced in the present work are contained in the manuscript.

## References

1. World Health Organization. World malaria report 2022. Geneva: WHO; 2022.

2. Snow RW, Guerra CA, Noor AM, Myint HY, Hay SI. The global distribution of clinical episodes of Plasmodium falciparum malaria. Nature. 2005;434(7030):214–7.

3. Egbom SE, Nduka FO, Nzeako SO. Point prevalence mapping of malaria infection in Rivers State, Nigeria. Tanzan J Health Res. 2022;23(4):1–8.

4. Menard D, Dondorp A. Antimalarial drug resistance: a threat to malaria elimination. Cold Spring Harb Perspect Med. 2017;7(7):a025619.

5. Zakai NA, Katz R, Hirsch C, et al. A prospective study of anemia status, hemoglobin concentration, and mortality in an elderly cohort. Arch Intern Med. 2005;165(19):2214–20.

6. Bousema T, Okell L, Felger I, et al. Asymptomatic malaria infections: detectability, transmissibility, and public health relevance. Nat Rev Microbiol. 2014;12(12):833–40.

7. Douglas NM, Anstey NM, Buffet PA, et al. The anaemia of Plasmodium vivax malaria. Malar J. 2012;11:135.

8. Nweneka CV, Doherty CP, Cox S, Prentice A. Iron delocalisation in the pathogenesis of malarial anaemia. Trans R Soc Trop Med Hyg. 2010;104(3):175–84.

9. Phillips RE, Pasvol G. Anaemia of Plasmodium falciparum malaria. Baillieres Clin Haematol. 1992;5(2):315–30.

10. Eziefula AC, Gosling R, Hwang J, et al. Rationale for short-course primaquine in Africa to interrupt malaria transmission. Malar J. 2012;11:360.

11. Echiejini CC. Effect of malaria on the reticulocyte count of female Madonna University students. [Unpublished dissertation]. Madonna University; 2024.

12. Egbom SE, Nduka FO, Nzeako SO. Malaria prevalence and hematological indices among university students in southern Nigeria. J Infect Dev Ctries. 2023;17(3):321–8.

13. Nigeria Malaria Indicator Survey 2021. National Malaria Elimination Programme; 2022.

14. Kish L. Survey sampling. New York: John Wiley & Sons; 1965.

15. Egbom SE et al. J Infect Public Health. 2022;15(3):287–94.

16. WHO. Nutritional anaemias: tools for effective prevention. Geneva: World Health Organization; 2017.

17. Cheesbrough M. District laboratory practice in tropical countries. 2nd ed. Cambridge: CUP; 2006.

18. WHO. Malaria microscopy quality assurance manual. Geneva: World Health Organization; 2016.

19. SD BIOLINE Malaria Ag Pf/Pan® package insert. Standard Diagnostics; 2021.

20. Bain BJ. Blood cells: a practical guide. 5th ed. Oxford: Wiley-Blackwell; 2015.

21. Riley RS et al. Lab Med. 2001;32(7):368–73.

22. IBM Corp. IBM SPSS Statistics for Windows, Version 25.0. Armonk, NY: IBM Corp; 2017.

23. FMOH. National guidelines for malaria diagnosis and treatment. Abuja: Federal Ministry of Health; 2021.

24. National Malaria Elimination Programme. Nigeria Malaria Indicator Survey 2022. Abuja: NMEP; 2023.

25. Adebayo AM, Akindele AA, Balogun IO, Chukwuemeka VI. High malaria prevalence among Nigerian university students. Trop Med Health. 2021;49:42.

26. Okeke TA, Udeze AO, Bello IS, Okafor HU. Age patterns of malaria infection in Nigerian adults. J Infect Public Health. 2020;13(5):712–718.

27. Lamikanra AA, Brown D, Potocnik A, Casals-Pascual C, Langhorne J, Roberts DJ. Malaria and erythropoiesis. Front Immunol. 2021;12:649162.

28. Tefferi A, Hanson CA, Inwards DJ. Reticulocyte reference ranges. Blood. 2019;134(Suppl_1):3575.

29. Adegoke OV, Okafor PN, Adepoju OT. Nutritional status of Nigerian students. J Nutr Sci. 2022;11:e35.

